# Evaluating Essential Coaching for Every Mother Tanzania (ECEM-TZ) as a postpartum text message digital health solution: A randomized controlled trial

**DOI:** 10.64898/2026.03.03.26347504

**Authors:** Justine Dol, Lilian Mselle, Marsha Campbell-Yeo, Columba Mbekenga, Douglas McMillan, Cindy-Lee Dennis, Gail Tomblin Murphy, Megan Aston

## Abstract

**Background:** Text messages are a low-cost digital health solution that can provide information directly to mothers. We aimed to evaluate a text message program, called *Essential Coaching for Every Mother Tanzania (ECEM-TZ),* designed to improve maternal access to essential newborn care education during the immediate 6-week postnatal period.

**Methods:** A randomized controlled trial was conducted in Dar es Salaam, Tanzania. E*CEM-TZ* consists of standardized text messages from birth to 6 weeks postpartum that provided evidence-based information on newborn care and recognizing danger signs. The primary outcome was newborn care knowledge. Secondary outcomes included parenting self-efficacy, breastfeeding self-efficacy, postpartum depression and anxiety symptoms, attendance at the six-week postnatal check-up, and newborn morbidity and mortality. Data were analyzed using ANCOVAs and logistical regression.

**Results:** Between June 13 and July 22, 2024, 143 mothers were randomized, 71 to the control group (standard care) and 72 to the intervention group (standard care plus *ECEM-TZ*), of which 139 completed both the baseline and follow-up at 6-8 weeks postpartum. Compared to the control group, mothers who received *ECEM-TZ* had significantly higher newborn care knowledge scores (MD=2.92, p<0.001) and fewer postpartum depression symptoms (MD=-1.55, p<0.001). Mothers who received *ECEM-TZ* were also three times more likely to attend a postnatal visit than those in the control group (OR=3.15, 95%CI [1.29, 7.72]).

**Conclusion:** Text messaging, as a low-cost, accessible digital health solution, is an important asset to enhance education of mothers in low- and middle-income countries during the immediate 6-week postpartum period.

**Trial registration:** ClinicalTrials.gov (NCT05362305), submitted 22-April-2022.

## INTRODUCTION

In Tanzania, the neonatal mortality ratio is 20 per 1,000 live births [1] and the maternal mortality ratio is 104 per 100,000 live births [2]. Despite the high number of deaths that occur postnatally, mothers frequently receive little education about their newborn’s care before hospital discharge or during the immediate postpartum period due to health system challenges [3, 4]. This also results in insufficient knowledge regarding infant danger signs in the postpartum period, which is a significant barrier to timely and life-saving care [5]. New mothers desire information from reliable sources about how to best care for their newborn during the postnatal period [6], yet challenges in delivery and uptake of this information have been noted, particularly in low-and middle-income countries (LMICs) [3]. When mothers are educated on newborn danger signs, their knowledge increases and they are more likely to seek appropriate care than those who did not receive such critical education [7]. There is a significant need to develop and evaluate innovative approaches to reach mothers during this period to ensure they are sufficiently knowledgeable and confident in caring for their newborns safely at home.

Mothers in LMICs also struggle with mental health and adjustment during the postpartum period. Approximately 1 in 5 mothers in Africa experience postpartum depression [8], and between 11% and 44% of mothers experience perinatal anxiety [9], both of which are a higher proportion than those in high-income countries [10]. Poor maternal mental health as been associated with low use of postnatal care services and negative child outcomes, including malnutrition, delays in immunization, and poor breastfeeding practices [10]. Parenting self-efficacy has been associated with reduced maternal mental health in the postpartum period, leading to improved child outcomes [11]. Thus, it is essential to address maternal psychosocial and mental health outcomes, particularly in LMICs, to improve not only maternal health but also infant and child well-being.

Digital health solutions can support mothers in the postpartum period by complementing existing care and addressing knowledge gaps by providing timely, evidence-based information and can be targeted to different contexts. Studies have found positive effects of digital health solutions on improving maternal awareness of newborn care and danger signs in the postpartum period, including in India [12], South Africa [13], and China [14]. The use of mobile phones to reach mothers is supported by the World Health Organization, who suggest that traditional postnatal care can be enhanced through mobile phone-based contact between mothers and the health system [15]. As such, our team developed a postpartum text message digital health solution called *Essential Coaching for Every Mother Tanzania (ECEM-TZ)* to provide evidence-based education directly to mothers to significantly enhance postnatal care for the infant and mother [16].

### Study Aims

To evaluate the effectiveness of *ECEM-TZ* as a postpartum mobile health solution at 6 weeks postpartum. We hypothesized that compared to standard care, mothers who received *ECEM-TZ* would have:

H1. Higher newborn care knowledge (*primary outcome*), parenting self-efficacy, and breastfeeding self-efficacy.

H2. Lower postpartum depression and postpartum anxiety symptoms.

H3. More frequent postnatal care attendance and lower newborn morbidity and mortality.

## METHODS

### Study Design

This study used a 2-arm, parallel-group, hybrid type 1 randomized controlled trial (RCT), following a pre-defined protocol [16]. This study was conducted in Dar es Salaam, Tanzania with ethics approval obtained from Dalhousie University (REB#2018-4538), Muhimbili University of Health and Allied Sciences (REB#MUHAS-REC-7-2019-010), and the National Medical Institute of Research (REB#NIMR/HQ/R.8c/Vol.I/2457). All procedures are in accordance with the Declaration of Helsinki. This paper reports on the effectiveness outcomes, with the implementation outcomes reported elsewhere.

### Participants

Mothers were eligible if they met the following criteria: (1) had given birth within the past 48 hours; (2) had daily access to a mobile phone, personal or shared, with text messaging capabilities; (3) were over 18 years of age; and (4) spoke and read English or Kiswahili. Participants were excluded if: (1) their newborn died or was expected to die before leaving the hospital; (2) they were unwilling or unable to receive text messages; (3) were experiencing major postnatal complications that were expected to impact learning capacity or ability to provide informed consent (e.g., postpartum haemorrhage, seizures) or (4) participated in *ECEM-TZ* development phase.

Mothers were recruited after giving birth at either the Mwananyamala Regional Referral Hospital or Sinza Palestina Hospital in Dar es Salaam, Tanzania. Mothers were approached by trained research assistants who visited the hospitals throughout the recruitment period and provided a detailed study explanation to potentially eligible mothers in the postpartum unit. Interested and eligible mothers provided informed consent and completed baseline data via Research Electronic Data Capture (REDCap) [17] before randomization. The research assistant collected data face-to-face for baseline with participants who then entered the data directly into the REDCap survey. Baseline data was collected June 13 and July 22, 2024 and the 6-week follow-up data collection occurred between July 30, and September 21, 2024.

### Randomization and Masking

Using a 1:1 allocation, participants were randomized into either the intervention group (standard care plus *ECEM-TZ*) or the control group (standard care) using a computer-generated stratified block randomization with blocks varying between 4 to 8 via REDCap. Stratification based on site of recruitment was implemented. Due to the complex and interactive nature of *ECEM-TZ*, the blinding of study participants was not possible. Mothers in the control group were not exposed to any *ECEM-TZ* text messages. To minimize bias at the participant level, all mothers received the same in-hospital care. Hospital staff (nurse midwives, physicians, and other healthcare providers) were blind to participant group allocation to ensure that both groups received standard discharge education. The research assistants involved in recruitment and data collection were not involved in the randomization process.

### Procedures

After completing informed consent procedures, participants completed a baseline questionnaire via a REDCap survey link facilitated by the research assistant. After baseline surveys were completed, participants were randomized to the intervention (*ECEM-TZ* plus standard care) or control (standard care). All participants received a phone voucher to their airtime provider of choice as an honorarium (10,000 Tanzanian shillings/∼ $5 Canadian). All participants were followed up by telephone by trained research assistants post intervention around 6 weeks postpartum with the research assistants entering the data directly into the REDCap survey.

#### Intervention

*ECEM-TZ* is a one-way text message program initiated on the third day after birth and provided until 6 weeks postnatally, to provide a total of 44 text messages. Information on the process of development of the messages is available in the protocol [16]. Messages were sent once per day in the morning and were available in Kiswahili, a native language of Tanzania. Messages pertained to the following topics: four on the research study, seven on infant feeding, six on postnatal care follow-up, four on normal infant development, three on infant thermal care, three on maternal care, two on cord care, and two on handwashing. For messages related to danger signs, three each were on fever and no movement/convulsions, two each were on poor feeding, jaundice, and breathing, and one was on cord infection. No other changes in care or education were implemented. While mothers could message “acha” (stop in Kiswahili) to withdraw from the study at any time and stop receiving messages, none did.

#### Control Group

Mothers in the control group did not receive any text messages from the program. There were no changes in standard postnatal care or education for the trial. All study participants received usual in-person education that was provided to all women delivering at that hospital before discharge from the postpartum unit.

### Sample Size

Sample size was calculated based on the primary outcome of newborn care knowledge where we considered a binary summative score of 100% the goal at 6 weeks postpartum. Using a power of 80%, alpha of .05 (two-tailed), and assuming 90% of the intervention group and 70% of the control group will score 100% at 6 weeks, a total sample size of 124 was required [18]. In ensure sufficient sample size, 12% attrition was accounted for with the total targeted sample size 141.

### Outcomes

#### Effectiveness Outcomes

##### Primary Outcome

Newborn care knowledge was assessed using a modified questionnaire developed by McConnell and colleagues, specifically related to newborn danger signs, handwashing practices, cord care, and newborn thermal care [19] where a total summative score was created (scores ranged from 0 to 15). Additionally, binary variables were created to determine whether mothers could identify three or more newborn danger signs, two or more best practice hand washing, two or more cord care practices, and three or more thermal care practices.[19] Based on these variables, a summative score was created ranging from 0 to 4, with an increased score representing more newborn care knowledge. This outcome was measured at baseline and 6 weeks postpartum.

##### Secondary outcomes

The following outcomes were measured at baseline and 6 weeks postpartum: parenting self-efficacy (Karitane Parenting Confidence Scale [20]), breastfeeding self-efficacy (Breastfeeding Self-Efficacy Scale – Short Form [21, 22]), postpartum depression (Edinburgh Postnatal Depression Scale (EPDS) [23, 24]), and postpartum anxiety (Generalized Anxiety Disorder (GAD)-7 [25, 26]). Additionally, 6-week postnatal clinic attendance (number attended, attend at least once), newborn mortality (dead/alive), and newborn morbidity (e.g., presence of at least one morbidity such as diarrhea, jaundice, infection) were collected.

### Statistical Analysis

Data was analyzed on an intention-to-treat basis using Statistical Package for Social Sciences version 29 (IBM SPSS Statistics, IBM Corporation, Somers, NY, USA). Demographic characteristics were expressed in means, standard deviations and percentages, as applicable, based on group allocation. Any significant differences in baseline characteristics, were examined through a chi-square analysis or t-test, as applicable, and were adjusted for in the analyses. A p-value of 0.05 was considered significant for all outcomes. For all analyses, unadjusted and adjusted models are reported. The trial is registered on ClinicalTrials.gov Identifier: NCT05362305.

For the effectiveness outcomes, total and summative scores are reported using means, standard deviations, and percentages, as appropriate. Analysis of Co-Variance (ANCOVA) was used to measure whether scores differed between the two groups, adjusted for the pre-test scores and any significant baseline characteristics. For newborn morbidity, mortality, and attendance at postnatal care six-week contact, binary variables were created, and data were analyzed using logistic regression. A t-test was used to determine any difference in the number of postnatal attendance visits.

## RESULTS

### Participants

A total of 173 participants were approached during recruitment between June 13 and July 22, 2024 (see Figure 1 for CONSORT flow). Of the 143 eligible participants who provided informed consent, 72 were randomized to the intervention group and 71 to the control group. Of this, 139 completed both the baseline and follow-up at 6-8 weeks postpartum, 68 in the intervention group and 71 in the control group.

**Figure 1.**
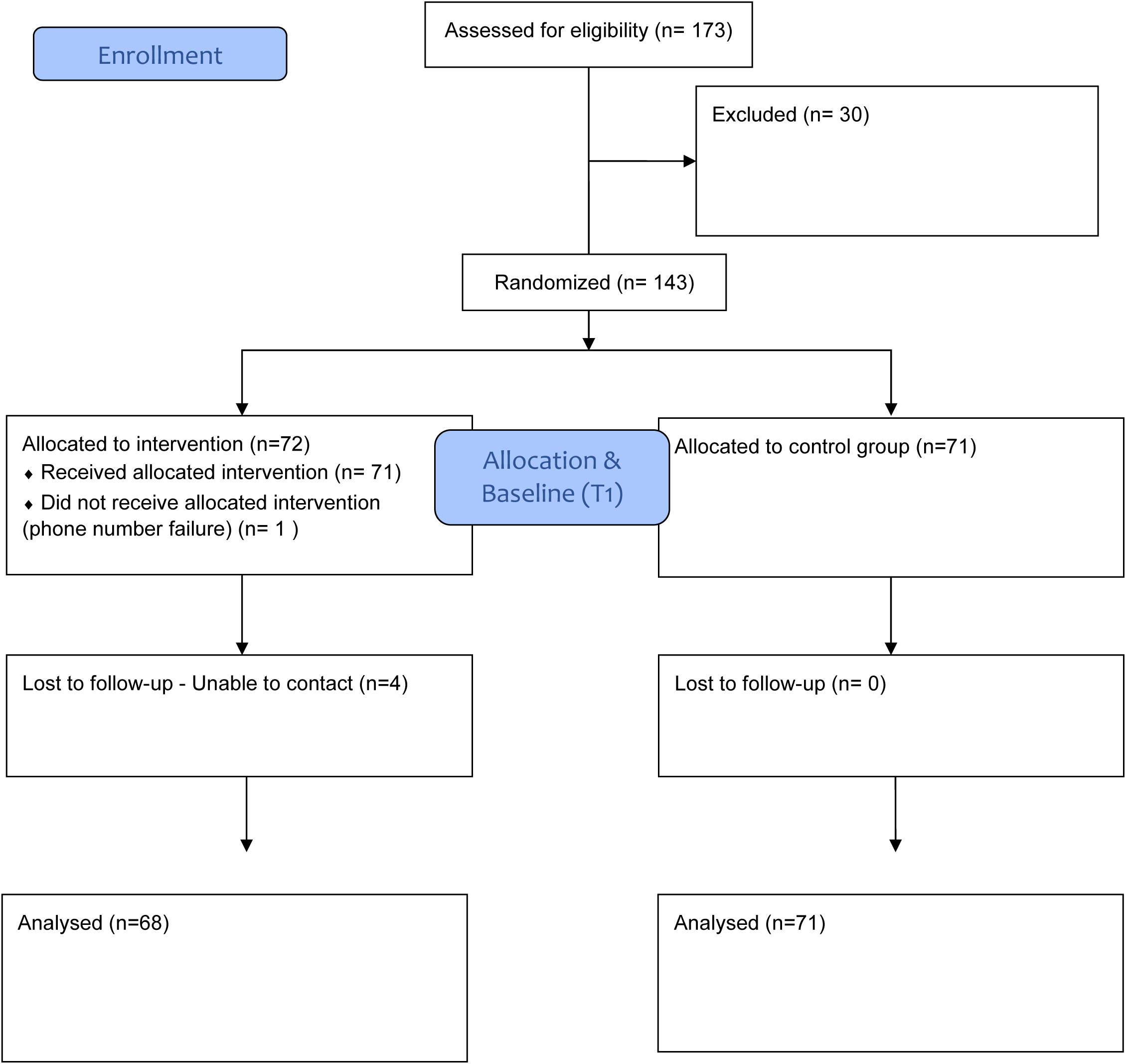
CONSORT Flow Diagram.

Mothers were on average 27.10 years (SD: 5.65) and 38.75 weeks gestation at birth (SD: 1.80). Sixty-three mothers were primiparous (44.1%) and the majority were married (67.8%). Table 1 describes baseline characteristics between the two groups. There was no significant difference between the groups on any of the primary or secondary outcomes at baseline (Table 2). Participants in the *ECEM-TZ* groups completed the follow-up evaluation on average 65 days (SD=9.8) postpartum compared to 58 (SD=8.81) days postpartum for those in the control group (p<0.001).

**Table 1.** Baseline characteristics of the intention-to-treat population.

|  | **ECEM-TZ (n=72)** | **Control (n=71)** |
| --- | --- | --- |
| Maternal Age (years) | 26.74 (5.03) | 27.48 (6.24) |
| Infant Age (days) | 0.56 (0.60) | 0.65 (0.59) |
| Child Sex |  |  |
| Male | 38 (52.8%) | 33 (46.5%) |
| Female | 34 (47.2%) | 38 (53.5%) |
| Parity |  |  |
| Primiparous | 34 (47.2%) | 29 (40.8%) |
| Multiparous | 38 (52.8%) | 42 (59.2%) |
| Type of birth |  |  |
| Vaginal | 48 (66.7%) | 53 (74.6%) |
| C-section | 24 (33.3%) | 18 (25.4%) |
| Marital Status |  |  |
| Single | 16 (22.2%) | 16 (22.5%) |
| Married | 47 (65.3%) | 50 (70.4%) |
| Other | 9 (12.5%) | 5 (7.1%) |
| Education |  |  |
| Standard 7 or below | 28 (38.9%) | 27 (38.0%) |
| Form 4 | 37 (51.4%) | 29 (40.8%) |
| Form 6 | 1 (1.6%) | 2 (2.8%) |
| University | 6 (8.3%) | 13 (18.3%) |

**Table 2.** Primary and secondary effectiveness outcomes among ECEM-TZ and control participants, unadjusted.

|  | **ECEM-TZ (n=72)** | **Control (n=71)** | **p-value** |
| --- | --- | --- | --- |
| **Baseline (pre-intervention)** |  |  |  |
| Newborn care knowledge (total) | 6.14 (2.36) | 5.94 (2.36) | .62 |
| Newborn care knowledge (binary) | 1.07 (1.13) | 1.07 (1.05) | .99 |
| Parenting self-efficacy | 36.67 (3.96) | 38.54 (4.15) | .10 |
| Breastfeeding self-efficacy | 62.27 (4.95) | 61.34 (6.04) | .33 |
| Postpartum depression symptoms | 9.33 (5.18) | 9.47 (5.47) | .88 |
| Postpartum anxiety symptoms | 8.42 (5.08) | 8.45 (5.55) | .97 |
|  | **ECEM-TZ (n=68)** | **Control (n=71)** | **p-value** |
| **6-Week (post-intervention)** |  |  |  |
| Newborn care knowledge (total) | 10.77 (2.41) | 7.85 (2.70) | <.001 |
| Newborn care knowledge (binary) | 3.34 (0.92) | 2.39 (1.05) | <.001 |
| Parenting self-efficacy | 40.54 (3.78) | 40.90 (3.34) | .091 |
| Breastfeeding self-efficacy | 63.83 (7.13) | 64.27 (6.09) | .555 |
| Postpartum depression symptoms | 3.63 (2.85) | 5.17 (2.65) | .001 |
| Postpartum anxiety symptoms | 1.42 (1.76) | 1.86 (1.89) | .147 |

### Outcomes

In the unadjusted analysis, mothers in the intervention group had significantly higher newborn care knowledge total score (M=10.76, SD=2.40) at 6 weeks postpartum compared to those in the control group (M=7.85, SD=2.70, p <0.001, Table 2). This was consistent for binary scores, where mothers in the intervention group had a significantly higher newborn care knowledge binary score (M=3.34, SD=0.92) at 6 weeks postpartum compared to those in the control group (M=2.39, SD=1.05), p <0.001. Additionally, mothers in the intervention group had fewer depression symptoms (M=3.63, SD=2.85) at 6 weeks postpartum than those in the control group (M=5.17, SD=2.65), p=0.001. In the unadjusted analysis, no other outcomes were significant.

Adjusting for parity and number of days postpartum, mothers in the intervention group continued to have significantly higher newborn care knowledge total score (M=10.73, SD=2.40) at 6 weeks postpartum compared to those in the control group (M=7.88, SD=2.70, p <0.001, Table 3). Additionally, adjusting for martial status, gestational age at birth, parity, and number of days postpartum, mothers in the intervention group had fewer depression symptoms (M=3.60, SD=2.85) at 6 weeks postpartum than those in the control group (M=5.20, SD=2.65), p=0.003.

**Table 3.** Primary and secondary effectiveness outcomes among ECEM-TZ and control participants, adjusted significantly correlated outcomes at baseline and relevant variables.

|  | **ECEM-TZ**<br>**(n=68)**<br>**M (SD)** | **Control**<br>**(n=71)**<br>**M (SD)** | **Mean**<br>**Difference** | **p-value** |
| --- | --- | --- | --- | --- |
| **6-Week (post-intervention)** |  |  |  |  |
| Newborn care knowledge <sup>a</sup> | 10.73 (2.40) | 7.88 (2.70) | 2.84 | <.001 |
| Parenting self-efficacy <sup>b</sup> | 40.08 (3.78) | 41.33 (3.34) | -1.25 | .028 |
| Breastfeeding self-efficacy <sup>a</sup> | 63.75 (7.13) | 64.35 (6.09) | -0.60 | .617 |
| Postpartum depression symptoms <sup>c</sup> | 3.60 (2.85) | 5.20 (2.65) | -1.60 | .003 |
| Postpartum anxiety symptoms <sup>d</sup> | 1.29 (1.76) | 1.98 (1.89) | -0.69 | .04 |

<sup>a</sup>controlled for parity and number of days postpartum at 6-week follow-up <sup>b</sup>controlled for gestation at birth, parity, and number of days
postpartum at 6-week follow-up <sup>c</sup>Controlled for marital status, gestation at birth, parity, and number of days postpartum at 6-week follow-up <sup>d</sup>

Furthermore, parenting self-efficacy was significantly different in the adjusted analysis, controlling for parity, gestation at birth, and number of days postpartum at follow-up. Mothers in the control group had higher parenting self-efficacy at 6 weeks postpartum (M=41.33, SD=3.34) compared to mothers in the intervention group (M=40.08, SD=3.78, p=0.028), yet both groups scored within the high parenting self-efficacy classification (≥40, Table 3). Mothers in the *ECEM-TZ* group had significantly lower postpartum anxiety scores (M=1.29, SD=1.76) than mothers in the control group (M=1.89, SD=1.89, p=0.04) when controlling for education, gestation at birth, parity, and number of days postpartum at 6-week follow-up.

No infants died during the trial period, so no analysis was possible on neonatal mortality. In relation to newborn morbidity, 18 mothers indicated their infant experienced at least one morbidity (i.e., fever, difficulty breathing, cord infection, lethargic, eye infection, convulsions, jaundice, diarrhea, poor feeding, problems urinating, or other), indicating a prevalence rate of 12.9%. There was no significant difference in likelihood of neonatal morbidity based on group allocation (OR=2.14, 95%CI [0.753, 6.07], p=0.153).

A logistic regression was performed to ascertain the effects of group allocation on the likelihood that mothers attended at least one postnatal clinic visit. The model was statistically significant, χ2(1)=6.89, p=0.009 and explained 75.0% (Nagelkerke R^2^) of the variance in postnatal clinic attendance and correctly classified 79.1% of cases. Mothers (and babies) in the intervention group were three times more likely to have at least one postnatal visit than control mothers (OR=3.15, 95%CI [1.29, 7.72]). Furthermore, in comparing the number of postnatal clinic contacts, mothers in the intervention group had significantly more postnatal visits (M=1.83, SD=0.94) than those in the control group (M=1.44, SD=0.76, p=0.019).

## DISCUSSION

This study evaluated the effectiveness of *ECEM-TZ* as a postpartum text message digital health solution to 6 weeks postpartum. Findings suggest that the *ECEM-TZ* intervention significantly improved newborn care knowledge and reduced postpartum depression and anxiety symptoms. In addition, mothers who received *ECEM-TZ* were significantly more likely to attend at least one postnatal clinic visit and overall completed more visits. Mothers in the *ECEM-TZ* group had higher parenting self-efficacy scores than mothers in the control group with both groups within the high parenting self-efficacy category. There was no difference in breastfeeding self-efficacy or newborn morbidity.

While previous text messaging digital health solutions have been evaluated in Tanzania (*Wazazi Nipendeni* (Love me, Parents) [27] and *Wired Mothers* [28]), they were developed and evaluated over a decade ago (2012-2014). Text messaging has been acknowledged as one of the potential digital health solutions that could be effectively used to significantly improve maternal, child, and neonatal health globally. Evidence linking newborn care knowledge for mothers in the perinatal period to newborn outcome is mixed [29]. By utilizing a low-cost, brief, co-designed text message program that focuses specifically on the critical first 6 weeks postpartum period, our findings suggest that text messages can improve maternal newborn care knowledge. More research is required to understand the contributing factors within digital health solutions that lead to an increased uptake and maintenance of knowledge among postpartum mothers.

This increase in newborn care knowledge, when combined with the finding that mothers who received *ECEM-TZ* were not only more likely to have at least one postnatal clinic visit but also more likely to participate in more visits, is promising. The utility of mobile health interventions, including text messaging, to improve postnatal care attendance is well established [29, 30]. Previous studies in Tanzania have attributed higher utilization of postnatal care to higher maternal education [31, 32], living in an urban area [31–33], receiving counselling from a healthcare provider to attend [31–33], higher socioeconomic status [32, 33], and having trust in the health system [31]. In this study, there were no differences between the groups based on education and all participants were recruited in an urban area. Thus, though the combined messages related to newborn care danger signs and counseling mothers to attend postnatal visits, this approach could have enhanced trust in the health system by providing evidence-based information.

*ECEM-TZ* was associated with lower postpartum depression and anxiety symptoms and higher parenting self-efficacy in the adjusted model. In a systematic review on mobile interventions in LMICs, maternal mental health and parenting self-efficacy was noted as severely underrepresented [29]. Given approximately 1 in 5 mothers in Sub-Saharan Africa experience postpartum depression [8], and that *ECEM-TZ* was able to reduce postpartum depression and anxiety symptoms in our trial, this finding is clinically important and consideration should be given to further expanded use. There is a general dearth of evidence on parenting self-efficacy and breastfeeding -efficacy in LMICs and even less in Africa [29]. This is an area ripe for future exploration to determine whether the measures used to identify parenting self-efficacy are culturally relevant to parents in LMICs or whether their self-efficacy related to parenting focuses on different outcomes, such as providing children with basic needs, having well-behaved children, and having good relationships with children [34].

### Strengths and Limitations

While this study was conducted with rigour, some limitations must be acknowledged. In addition to the limitations reported in the protocol [16], the primary limitation is that there was significant difference in the number of days postpartum between the intervention and control group. This resulted in the intervention group being followed up on average a week later than the mothers in the control group. However, controlling for this in the adjusted analysis did not substantially impact most outcomes. Another limitation is that we only collected data immediately post-intervention and cannot report on any longer-term follow-up impacts.

### Future Directions

As this study recruited mothers solely from Dar es Salaam, the commercial capital of Tanzania, exploring potential expansion in non-urban areas and scaling to other countries will be important to examine generalizability. Additionally, there are opportunities to partner with other digital health solutions, such as Afya Mama, which is an offline informational mobile application that has been developed [35], but has not yet been evaluated for implementation in Tanzania. There is also the potential to utilize emerging technologies, such as artificial intelligence, to expand the reach of digital health solutions globally. Future research should also continue to explore the role of parenting self-efficacy and breastfeeding self-efficacy in low- and middle-income country contexts.

### Conclusions

Mothers who received *ECEM-TZ* had higher newborn care knowledge and parenting self-efficacy and lower postpartum depression and anxiety symptoms than mothers who did not. In addition, mothers who received *ECEM-TZ* were three times as likely to attend a postnatal visit as control mothers. This suggests that text messaging during the postpartum period can be a low-cost, accessible digital health solution to support mothers in low- and middle-income countries.

## Declarations

### Ethics approval and consent to participate

Ethics approval was obtained from Dalhousie University (REB#2018-4538), Muhimbili University of Health and Allied Sciences (REB#MUHAS-REC-7-2019-010), and the National Medical Institute of Research (REB#NIMR/HQ/R.8c/Vol.I/2457).

### Consent for publication

Not applicable.

### Availability of data and materials

The datasets used and/or analysed during the current study are available from the corresponding author on reasonable request.

## Acknowledgements

The authors would like to thank the research assistants (Vaileth Noel Pallangyo, Emmanuel Peter Massawe, Restituta Thadeus Mushi, Highness Mlay, Agape Bhoke) for their help with data collection throughout this project, as well as the mothers who participated in the development of messages and the study conduct.

## Author Contributions

JD: conceptualization, methodology, writing – original draft, writing – review and editing, project administration, funding acquisition

MCY: conceptualization, methodology, writing – review and editing, project administration, funding acquisition, supervision

LM: conceptualization, methodology, writing – review and editing, project administration CM: conceptualization, methodology, writing – review and editing, project administration, funding acquisition

DM: conceptualization, methodology, writing – review and editing C-LD: methodology, writing – review and editing

GTM: conceptualization, methodology, writing – review and editing MA: conceptualization, methodology, writing – review and editing

## References

1. World Bank. Mortality rate, neonatal (per 1,000 live births) - Tanzania. World Bank Open Data. https://data.worldbank.org. Accessed 22 Sep 2023.

2. Ministry of Health, Community Development, Gender, Elderly and Children (MoHCDGEC) [Tanzania Mainland], Ministry of Health (moH) [Zanzibar], National Bureau of Statistics (NBS), Office of the Chief Government Statistician (OCGS), ICF. Tanzania 2022 Demographic and Health Survey and Malaria Indicator Survey 2022 Summary Report. Dodoma, Tanzania and Rockville, Maryland, USA: MoHCDGEC, MoH, NBS, OCGS, and ICF; 2022.

3. Mselle LT, Aston M, Kohi TW, Mbekenga C, Macdonald D, White M, et al. The challenges of providing postpartum education in Dar es Salaam, Tanzania: Narratives of nurse-midwives and obstetricians. Qual Health Res. 2017;27:1792–803.

4. Dol J, Kohi T, Campbell-Yeo M, Tomblin Murphy G, Aston M, Mselle L. Exploring maternal postnatal newborn care postnatal discharge education in Dar es Salaam, Tanzania: Barriers, facilitators and opportunities. Midwifery. 2019;77:137–43.

5. Shannon K, Burridge J, Franklin B, Bhushan S, Hilsenbeck S, Petrova EV, et al. Gambian Mothers Lack Obstetric Danger Sign Knowledge, But Educational Intervention Shows Promise. Ann Glob Health. 2024;90:31.

6. Walker L, Murphey CL, Nichols F. The broken thread of health promotion and disease prevention for women during the postpartum period. J Perinat Educ. 2015;24:81–92.

7. Fujino Y, Sasaki S, Igarashi K, Tanabe N, Muleya CM, Tambatamba B, et al. Improvement in mothers’ immediate care-seeking behaviors for children’s danger signs through a community-based intervention in Lusaka, Zambia. Tohoku J Exp Med. 2009;217:73–85.

8. Nweke M, Ukwuoma M, Adiuku-Brown AC, Okemuo AJ, Ugwu PI, Nseka E. Burden of postpartum depression in sub-Saharan Africa: An updated systematic review. South Afr J Sci. 2024;120:1–12.

9. Abajobir A, Sidze EM, Wainaina C, Gerbaba MJ, Wekesah FM. The epidemiology of maternal mental health in Africa: a systematic review. Arch Womens Ment Health. 2025. 10.1007/s00737-025-01563-4.

10. Pan T, Zeng Y, Chai X, Wen Z, Tan X, Sun M. Global Prevalence of Perinatal Depression and Its Determinants Among Rural Women: A Systematic Review and Meta-Analysis. Depress Anxiety. 2024;2024:1882604.

11. Fang Y, Boelens M, Windhorst DA, Raat H, van Grieken A. Factors associated with parenting self-efficacy: A systematic review. J Adv Nurs. 2021;77:2641–61.

12. El Ayadi AM, Diamond-Smith NG, Duggal M, Singh P, Sharma P, Kaur J, et al. Preliminary impact of an mHealth education and social support intervention on maternal health knowledge and outcomes among postpartum mothers in Punjab, India. BMC Pregnancy Childbirth. 2025;25:239.

13. Coleman J, Eriksen J, Black V, Thorson A, Hatcher A. A Qualitative User Study of a Maternal Text Message-based mHealth Intervention: MAMA South Africa (Preprint). JMIR Hum Factors. 2019;7.

14. Xie R-HH, Tan H, Taljaard M, Liao Y, Krewski D, Du Q, et al. The impact of a maternal education program through text messaging in rural china: Cluster randomized controlled trial. JMIR MHealth UHealth. 2018;6:1–10.

15. World Health Organization. Postnatal care of the mother and newborn 2013. World Health Organization. 2013;:1–72.

16. Dol J, Mselle LT, Campbell-Yeo M, Mbekenga C, Kohi T, McMillan D, et al. Essential Coaching for Every Mother Tanzania (ECEM-TZ): Protocol for a Type 1 Hybrid Effectiveness-Implementation Randomized Controlled Trial. JMIR Res Protoc. 2024;13:e63454.

17. Harris PA, Taylor R, Thielke R, Payne J, Gonzalez N, Conde JG. Research electronic data capture (REDCap) – A metadata-driven methodology and workflow process for providing translational research informatics support. J Biomed Inf. 2009;42:377–81.

18. Fralick Lab. Power and Sampe Size Calculator. 2024. www.powercalc.ca. Accessed 9 Oct 2024.

19. McConnell M, Ettenger A, Rothschild CW, Muigai F, Cohen J. Can a community health worker administered postnatal checklist increase health-seeking behaviors and knowledge?: Evidence from a randomized trial with a private maternity facility in Kiambu County, Kenya. BMC Pregnancy Childbirth. 2016;16:136.

20. Črnčec R, Barnett B, Matthey S. Development of an instrument to assess perceived self-efficacy in the parents of infants. Res Nurs Health. 2008;31:442–53.

21. Mituki DM, Tuitoek PJ, Varpolatai A, Taabu I. Translation and validation of the breast feeding self efficacy scale into the Kiswahili language in resource restricted setting in Thika – Kenya. Gjmedph. 2017;6:1–9.

22. Dennis C. The Breastfeeding Self-Efficacy Scale: Psychometric Assessment of the Short Form. J Obstet Gynecol Neonatal Nurs. 2003;32:734–44.

23. Linnet Ongeri MK. Translation of EPDS Questionnaire into iswahili: Understanding the Cross-Cultural and Translation Issues in Mental Health Research. J Pregnancy Child Health. 2015;02:1–10.

24. Cox J, Holden J, Sagovksy R. Detection of Postnatal Depression: Development of the 10-item Edinburgh Postnatal Depression Scale. Br J Psychiatry. 1987;150:782–6.

25. Nyongesa MK, Mwangi P, Koot HM, Cuijpers P, Newton CRJC, Abubakar A. The reliability, validity and factorial structure of the Swahili version of the 7-item generalized anxiety disorder scale (GAD-7) among adults living with HIV from Kilifi, Kenya. Ann Gen Psychiatry. 2020;19:1–10.

26. Spitzer RL, Kroenke K, Williams JBW, Löwe B. A brief measure for assessing generalized anxiety disorder: The GAD-7. Arch Intern Med. 2006;166:1092–7.

27. Kaufman MR, Harman JJ, Smelyanskaya M, Orkis J, Ainslie R. “Love me, parents!”: Impact evaluation of a national social and behavioral change communication campaign on maternal health outcomes in Tanzania. BMC Pregnancy Childbirth. 2017;17:1–10.

28. Lund S, Hemed M, Nielsen BB, Said A, Said K, Makungu MH, et al. Mobile phones as a health communication tool to improve skilled attendance at delivery in Zanzibar: A cluster-randomised controlled trial. BJOG Int J Obstet Gynaecol. 2012;119:1256–64.

29. Dol J, Campbell-Yeo M, Tomblin Murphy G, Aston M, Mcmillan DD, Richardson B. Impact of mobile health interventions during the perinatal period for mothers in low- and middle-income countries: a systematic review. JBI Database Syst Rev Implement Rep. 2019;17:1634–67.

30. Hailemariam T, Atnafu A, Gezie LD, Tilahun B. Effect of short message service reminders in improving optimal antenatal care, skilled birth attendance and postnatal care in low-and middle-income countries: a systematic review and meta-analysis. BMC Med Inform Decis Mak. 2024;25:1.

31. Mohan D, Gupta S, LeFevre A, Bazant E, Killewo J, Baqui AH. Determinants of postnatal care use at health facilities in rural Tanzania: multilevel analysis of a household survey. BMC Pregnancy Childbirth. 2015;15:282.

32. Mahiti GR, Mkoka DA, Kiwara AD, Mbekenga CK, Hurtig AK, Goicolea I. Women’s perceptions of antenatal, delivery, and postpartum services in rural Tanzania. Glob Health Action. 2015;8:28567.

33. Larsen A, Exavery A, Phillips JF, Tani K, Kanté AM. Predictors of health care seeking behavior during pregnancy, delivery, and the postnatal period in rural Tanzania. Matern Child Health J. 2016;20:1726–34.

34. Augustinavicius JL, Familiar-Lopez I, Winch PJ, Murray SM, Ojuka C, Boivin MJ, et al. Parenting self-efficacy in the context of poverty and HIV in Eastern Uganda: A qualitative study. Infant Ment Health J Infancy Early Child. 2019;40:422–38.

35. Kullian NC, Ally JS, Magemo A. Mobile Health Application to Strengthen Postnatal Care: A Case of Tanzania. East Afr J Inf Technol. 2023;6:15–33.

